# Metagenomic analysis of common intestinal diseases reveals relationships among microbial signatures and powers multi-disease diagnostic models

**DOI:** 10.1101/19013136

**Authors:** Puzi Jiang, Sicheng Wu, Qibin Luo, Xing-ming Zhao, Wei-Hua Chen

## Abstract

Common intestinal diseases such as Crohn’s disease (CD), ulcerative colitis (UC) and colorectal cancer (CRC), share clinical symptoms and altered gut microbes, necessitating cross-disease comparisons and the use of multi-disease models. Here, we performed meta-analyses on thirteen fecal metagenome datasets of the three diseases. We identified 87 species and 65 pathway markers that were consistently changed in multiple datasets of the same diseases. According to their overall trends, we grouped the disease-enriched marker species into disease-specific and -common clusters, and revealed their distinct phylogenetic relationships: species in CD-specific cluster are phylogenetically related, while those in CRC-specific cluster are more distant; strikingly, UC-specific species are phylogenetically closer to CRC, likely because UC-patients have higher risk of CRC. Consistent to their phylogenetic relationships, marker species had similar within-cluster and different between-cluster metabolic preferences. There were part of marker species and pathways correlated with an indicator of leaky gut, suggesting a link between gut dysbiosis and human derived contents. Marker species showed more coordinated changes and tighter inner-connections in cases than the controls, suggesting that the diseased gut may represent a stressed environment and pose stronger selection to gut microbes. With the marker species and pathways, we constructed four high-performance (including multi-disease) models with AUROC of 0.87 and true positive rates up to 90%, and explained their putative clinical applications. We identified consistent microbial alterations in common intestinal diseases, revealed metabolic capacities and the relationships among marker bacteria in distinct states, and supported the feasibility of metagenome-derived multi-disease diagnosis.

**Importance:** Gut microbes have been identified as potential markers in distinguishing patients from controls in colorectal cancer, ulcerative colitis and Crohn’s disease individually, whereas there lacks a systematic analysis to investigate the exclusive microbial shifts of these enteropathies with similar clinical symptoms. Our meta-analysis and cross-disease comparisons identified consistent microbial alterations in each enteropathy, revealed microbial ecosystems among marker bacteria in distinct states, and demonstrated the necessity and feasibility of metagenome-based multi-disease classifications. To the best of our knowledge, this is the first study that constructed multi-class models in these common intestinal diseases.

## Background

In recent years, the incidences of several intestinal diseases including inflammatory bowel disease (IBD) and colorectal cancer (CRC) have been increasing in developmental countries while remained high in major western countries, mostly due to industrial urbanization and Western life-styles (1-6). For example, IBD, comprising mainly Crohn’s disease (CD) and ulcerative colitis (UC), has increased incidence in newly industrialized countries in Africa, Asia and South America (7); populations previously considered ‘low risk’ including Indian and Japanese also witnessed significant increase in incidence (6). In addition, as the overall incidence of CRC remained high in major western countries, an alarming trend of increased risk has been observed in young adults (3, 4).

IBD and CRC share several symptoms, including rectal bleeding, abdominal pain, diarrhoea, weight loss, and anaemia; furthermore, CRC in young patients has similar ages of onset to IBD (<50 years) (8). In addition, patients with IBD are considered at high risk of developing colorectal cancer, due to duration of inflammation and expansion of lesions. The accumulative risk of CRC in IBD patients is increasing over time (9). The European Crohn’s and Colitis Organisation (ECCO) and the American Gastroenterological Association (AGA) recommend that the IBD patients need to strengthen CRC surveillance with colonoscopies. But long-period surveillance doesn’t solve the problem because of deficiencies of regular colonoscopies in detecting dysplasia and other high risk factors in the elderly patients (10). It thus can be challenging to accurately separate these diseases in clinical practice, especially in their early stages and/or in younger patients; delay in diagnosis is common and can cause harm, as a recent research has pointed out (8).

Recent studies suggested that IBD and CRC are linked with complicated interplay of various components, involving genetics, environmental factors, gut microbiome and immune system (11, 12). So far, a few hundreds of genes have been identified, whose mutations and/or dysregulation of expressions were linked to increase risk of IBD and CRC (13-16). However, genetic factors together can only explain limited proportions of the disease incidences (17-19). Conversely, other factors are believed to be major contributors, especially gut microbes (12, 20-22). The latter, along with metabolites and antibiotics that they produced through digesting nutrients from food, the host and other microbes, could play important roles in modulating host immunity and inflammation, maintaining gastrointestinal equilibrium and resisting alien invaders (23).

Fecal microbial dysbiosis in IBD and CRC have been observed, and subsequently utilized to generate predictive models for patient stratification and/or risk evaluation (22, 24-26). For example, IBD patients showed a reduction of taxa from Firmicutes phylum and enrichment of pathogenic species (25, 27). Several studies showed the IBD subtypes, namely CD and UC had distinctive gut microbiota and metabolic profiles, though results differ across studies (28). Gut microbes have been identified as potential markers in distinguishing patients from controls in IBD and CRC individually, as both the increase of pathogens and development of lesions in gut contribute to the dysbiosis through affecting metabolic functions of bacteria (24, 29, 30).

However, binary models (i.e. models capable of distinguishing patients of a particular disease from controls) created for a single disease may lead to misdiagnosis, on account of some microbes commonly changed in diseases (31). Furthermore, most models, especially those available for IBD and/or its subtypes, were generated on data from a single population and may not perform well on other populations (25, 27). In addition, though limited by the use of 16S amplicon sequencing data with low resolution, a study across multiple diseases to search for disease-specific markers raised the issue that whether we could distinguish one gut illness from others using solely the gut microbiome data (31).

In sum, it is necessary to perform cross-disease comparisons and generate multi-class models capable of distinguishing these common intestinal diseases which can have very similar symptoms and associate with consistent gut microbiome alterations. It is also necessary to perform meta-analysis to account for population-specific biases. Meta-analysis is a method combining diverse projects and help us avoid biases from individual study (32); moreover, the latest surveys about CRC via meta-analysis suggest the necessity in collecting metagenome data as much as possible to identify consistently altered microbes in certain state (33, 34).

In this research, we collected thirteen metagenomics datasets for common intestinal diseases known to have strong links to gut microbiota, including three, three, and seven datasets for CD, UC and CRC respectively, and performed meta-analysis to (1) determine disease-specific and consistent microbial alterations, (2) elucidate possible mechanisms underlying the altered species associated with different disease states, and (3) generate high-performance multi-class models using taxonomic and metabolic profiles for easier and better clinical applications.

## RESULTS

### Collection and annotation of thirteen gut metagenomics datasets for common intestinal diseases

To determine consistently altered gut microbial features in common intestinal diseases such as UC, CD and CRC as comparing with controls, we conducted a systematic search in public databases (Fig. S1) and collected in total thirteen metagenomic datasets, including three, three and seven datasets for UC, CD and CRC respectively, totaling 763 cases and 632 controls (Table S1). MetaPhlAn2 and HUMAnN2 were used to determine the taxonomic and functional profiles of all samples.

Taxonomic analysis revealed that the alpha diversity was not significantly changed in all but one CRC datasets (PRJDB4176) as compared with their respective controls; conversely, alpha diversity was decreased significantly in patients of most CD datasets, while did not show consistent trends in UC patients (Table S1, Wilcoxon rank sum test, P-value < 0.05). Interestingly, we found that the human DNA contents (HDCs), as calculated as the percentage of sequencing reads mapped to the human genome, were significantly higher in patients in all diseases except one UC dataset (PRJEB1220) (Table S1), consistent with our results (35) that HDC could be used as a marker for intestinal diseases; the increased level of HDCs are likely due to that high level of deciduous epithelial and/or blood cells could be found in stools of patients with IBD or CRC resulting from gut injury and quickening cell cycles (21, 24, 29, 36).

### Disease-specific and shared taxonomic gut-microbiome markers in CRC, UC and CD

We used MaAsLin2, a multivariable analysis tool, on relative abundance of species to adjust the confounding factors, such as body mass index (BMI), gender and age, and identify differential species in each dataset. We then performed meta-analysis on each disease to identify microbes that showed consistent trends in the same disease and referred them as “marker species” accordingly. Consequently, we identified in total 14, 43 and 44 marker species in UC, CD and CRC respectively; among which, 8 (57.1% out of 14), 32 (74.4%) and 31 (70%) were unique to the respective diseases. Out of the in total 87 marker species, 14 were found in at least two diseases and no one was common to all diseases (Fig. 1A).

**Fig. 1.**
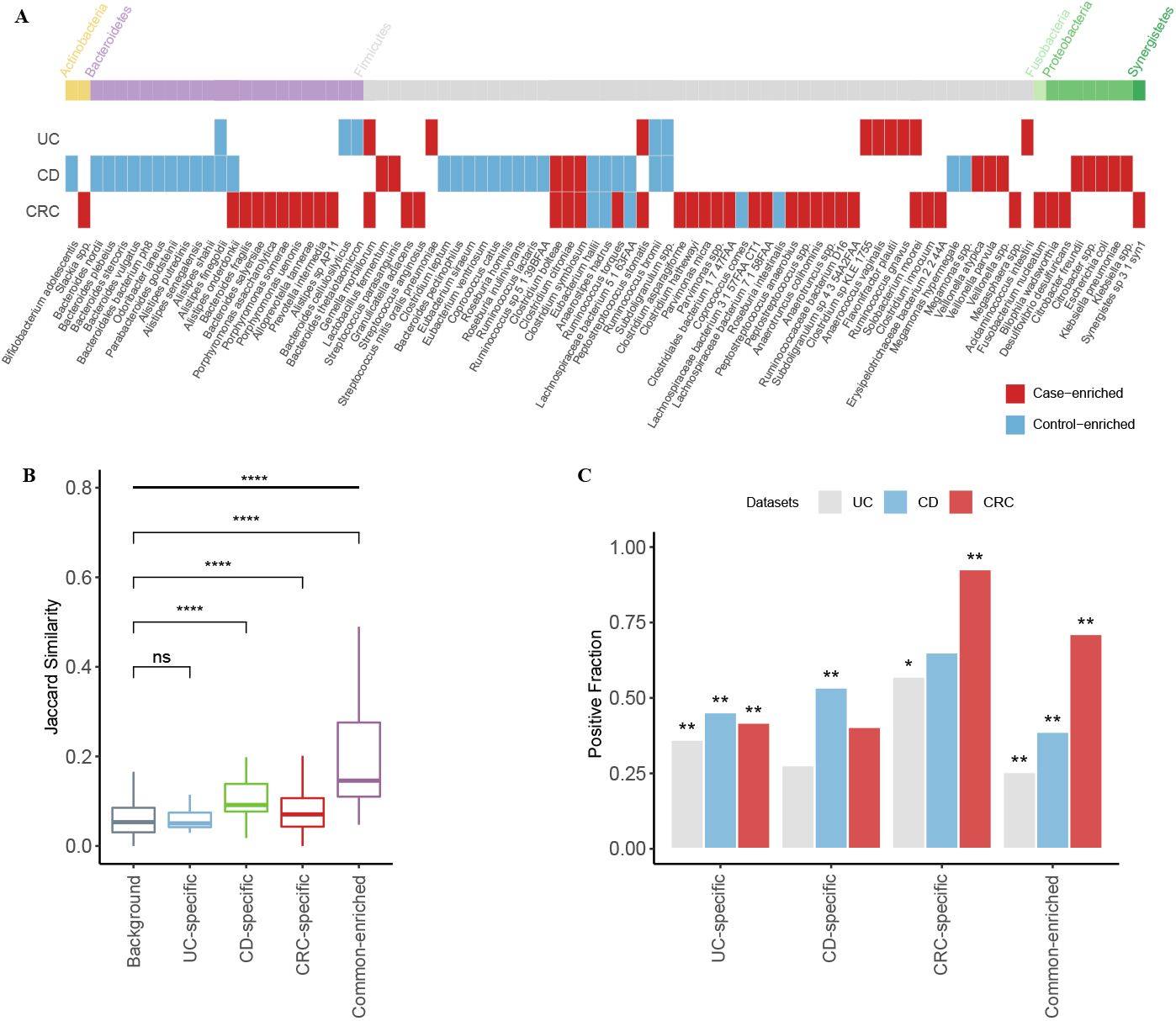
Disease-specific and shared microbial markers showed distinct prevalence profiles in patients and controls. **A.** Microbial markers and their trends (i.e. case- or control-enriched) in common intestinal diseases. Species significantly enriched in cases (or controls) of corresponding disease in meta-analysis are shown (fdr < 0.05, Benjamini-Hochberg FDR correction), with their phylum shown on top. Red indicates case-enriched species, and blue indicates control-enriched one. **B.** The boxplot shows the inner Jaccard similarities of case-enriched microbes in all cases. The case-enriched microbes were clustered according to their trends in intestinal diseases (see Materials and Methods). The background means the similarities between members that did not belong to the same cluster. The four clusters were named according to their members; UC-specific, CD-specific and CRC-specific clusters only contained the disease-specific markers, while common-enriched cluster contained markers from at least two diseases (****: p < 0.0001). **C.** The barplot shows the fraction of cases that are “positive” for given clusters in per disease type. Here positive samples for a given species are defined as those in which the species was found with higher relative abundance than 95% of all controls. The significant differences of the positive fraction between controls and cases for each cluster were assessed via Crochran-Mantel-Haenszel test with ‘dataset’ as the stratified factor; the ‘star’ symbols on the bars were used to indicate that marker species of a given cluster were significantly more prevalent in cases than in their corresponding controls (*: p < 0.05, **: p < 0.01).

For CRC, marker species were mostly disease-enriched, including *Fusobacterium nucleatum, Parvimonas micra, Gemella morbillorum and Peptostreptococcus stomatis*, most of which were reported widely (33, 34). Interestingly, a significant proportion of CRC-enriched marker species were identified significantly in majority of CRC datasets, while most of CRC-depleted marker species were dataset-specific (p-value < 0.05 identified by MaAsLin2, Fig. S2), which was also in accordance with previous studies (33, 34).

Conversely, CD patients showed a depletion of control-enriched species, including Roseburia inulinivorans, *Roseburia hominis, Coprococcus catus* and several members of genus *Alistipes, Bacteroides*, and *Eubacterium*, which were also consistent with previous studies (25, 27). However, marker species in UC were a mix of both but mainly driven by disease-enriched ones (Fig. 1A), unlike a recent study that primarily showed a decreasing of control-enriched species (27); the discrepancies are likely due to study-specific results (Fig. 1A). Moreover, most of UC- and CD-marker species were identified as significant differential species in at least two datasets, unlike the CRC marker species (p-value < 0.05 identified by MaAsLin2, Fig. S2).

Among the shared markers, *Alistipes onderdonkii* and *Ruminococcus torques* showed conflicted trends between disease; for example, they both were decreased in CD patients but increased in CRC patients. These results are in fact consistent with previous studies (37-40) and suggest that both overgrowth and loss of certain species represent the disturbance of intestinal environment.

Disease-enriched species are often directly linked to pathogenesis and direct targets for disease intervention. We thus first focused on these species and grouped them into disease-specific and common according to their shifts in intestinal diseases. As shown in Fig. 1A, in total 6, 10, 31 and 6 markers were assigned to UC-specific, CD-specific, CRC-specific and common-enriched groups respectively. Their phylogenetic relationships based on the NCBI taxonomic tree revealed their distinct distributions (Fig. S3). Firstly, CRC-specific markers whose taxonomic level were across six phyla, showed more diverse phylogenetically than CD- and UC-specific markers. CD-specific markers were members of two phyla, namely Proteobacteria and Firmicutes, including species within *Veillonella* genus, Enterobacteriaceae family, and Lactobacillales order. The UC-specific markers consist of species from Firmicutes. In addition, UC-specific species are phylogenetically closer to CRC-specific species, likely because UC patients have a little higher risk of CRC (9). Together, we revealed phylogenetic patterns of the marker microbes that could only be revealed through cross-disease analysis.

We next checked if the disease-enriched species could also show distinct prevalence patterns in their respective diseases vs. controls and/or in other diseases. To overcome the variances in species abundances, we defined a dynamic threshold for each species as its 95% quantile relative abundance of all control samples, and determined whether a species was present in a sample or absent (see Materials and Methods). By doing so, we obtained a binarized matrix with each row representing a disease-enriched species, and each column representing a patient. We calculated the Jaccard similarity to investigate the co-occurrence patterns of the clustered groups in all patients; as shown in Fig. 1B, we found that the markers in the CD-specific, CRC-specific, and common groups showed significantly higher inner-similarity in patients, indicating these species were preferably to co-exist with the members in the same group; however, the UC-specific species did not display such a trend.

We then summed up the prevalence of the disease-enriched species of each sample. As expected, these disease-specific markers were significantly enriched in their respective diseases, as well these common microbes were significantly enriched in all diseases (Fig. 1C). The UC-specific species, though lacked a strong co-occurrence among themselves, had a remarkable prevalence in each disease dataset. The results suggest that with the disease-specific clusters, it would be possible to stratify different diseases using microbial profiles, while the shared enriched species increased difficulties of classification.

### Disease-specific and shared functional markers in CRC, UC and CD

Using the same criteria, we identified in total 10, 37 and 39 marker pathways for UC, CD and CRC (Fig. S4); among which, 3 (30% out of 10), 18 (48.6%) and 25 (64.1%) were unique to these diseases respectively. Most of the UC and CD marker pathways were control-enriched and associated with biosynthesis, consistent to previous results (27). For example, pathways for amino acid biosynthesis, such as L-methionine biosynthesis I, and aspartate superpathway were depleted in CD patients, indicating the nutrient transport and uptake (28, 41). Conversely, the gut community in CRC showed distinct characteristics with the decreased capacities of carbohydrate degradation, and increased capacities on amino acid degradation, which were accordance with previous studies (33, 34).

To provide an overview on the changed functional capacities of gut microbes, we summarized the metabolic functions as the modules according to their superclasses in MetaCyc database (42). We applied the differential abundance analysis in module level, and found the characteristic functional pattern of IBD and CRC (Fig. 2). As mentioned previously, the module of amino acid degradation was decreased in CD patients, while its trend behaved in the opposite way in CRC patients. In CD patients, the elevated module about nucleoside and nucleotide degradation, which was comprised of degradation of purine, would induce the gut metabolic stress and involve in the inflammatory processes (43, 44). As essential pathways for energetic and biosynthetic demands of cancer cells, the carbohydrate biosynthesis, fermentation and glycolysis were enhanced in CRC (45, 46). In addition, as subtypes of IBD, CD and UC had distinct preferences in module level, though there was a high-degree overlapping among their associated pathways (Fig. S4).

**Fig. 2.**
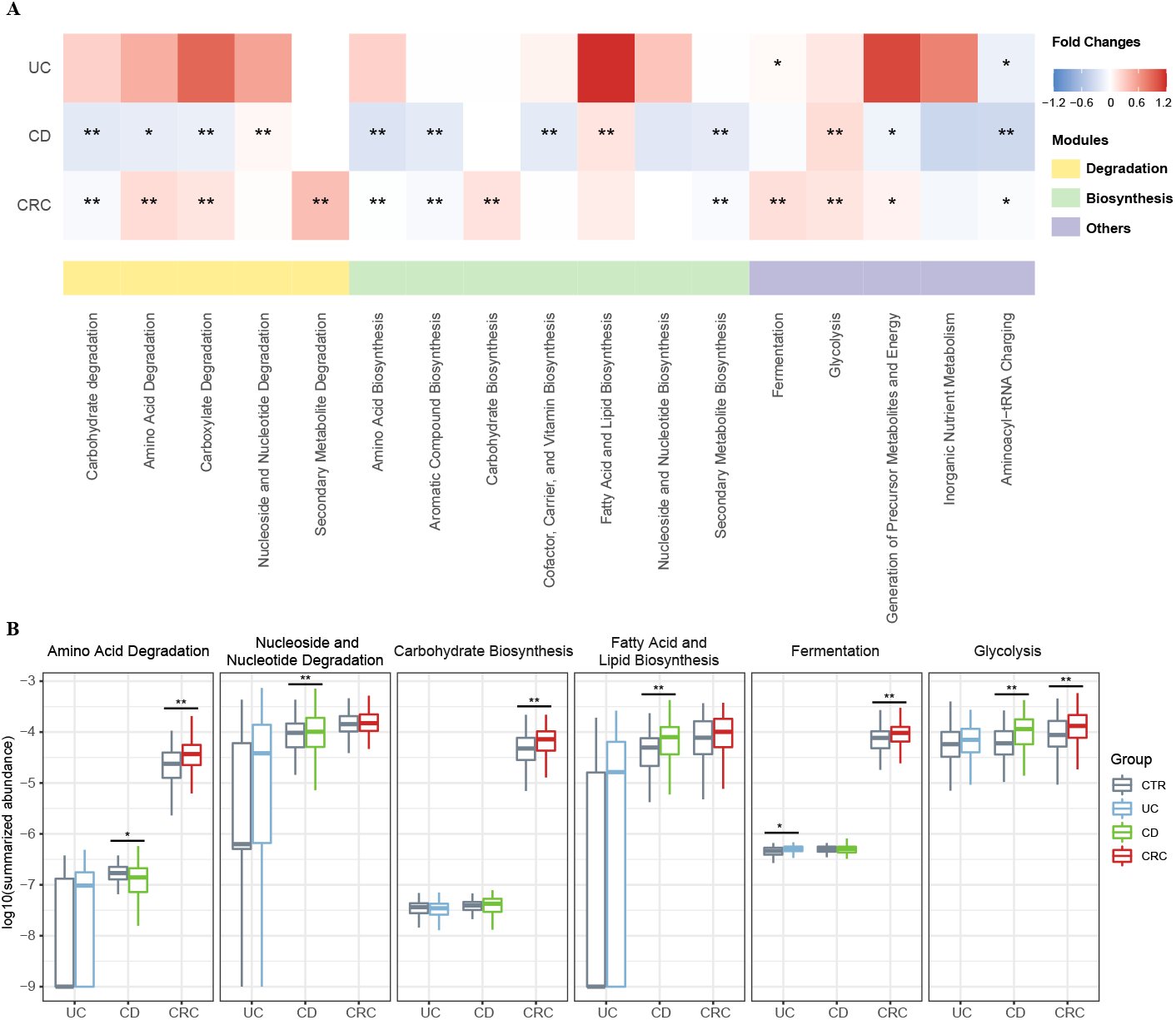
Disease-specific pattern of metagenomics functional modules. **A.** Functional modules were summed according to the category of MetaCyc database. The differences between controls and cases in one specified disease were calculated as the generalized fold changes, and the significances were assessed using two-sided Wilcoxon rank sum test and blocked with “datasets” (see Materials and Methods). Red indicates case-enriched modules, and blue indicates control-enriched modules. The asterisk indicated the module were significantly different between cases and controls (*: p < 0.05, **: p < 0.01). **B.** The boxplots show the distribution of some representative modules in the three intestinal diseases. The asterisks on the bars were calculated as mentioned above, indicating the significances.

Together, we identified consistently altered marker species and functional pathways in each of the intestinal diseases. A significant proportion of them were shared by two diseases while majority of them remained disease-specific. Of note, UC was associated with the least number of disease markers and the least proportion of unique ones.

### Disease marker microbes underlie altered metabolic capacities, especially in degradation

To check if the altered marker species could underlie the changes in metabolic capacities, we calculated partial Spearman’s rank correlations between marker species and metabolic pathways and performed meta-analysis to aggregate coefficients. Interestingly, we found that most of the disease-altered pathways showed statistically correlations with the marker species; more importantly, we were able to recaptulate the species clusters (including the control-enriched cluster identified in the previous sections) using their correlated metabolic capacities, especially in degradation (Fig. 3 &Fig. S5). For example, most of control-enriched species in CD and CRC, including members of genus *Coprococcus, Roseburia, Ruminococcus* and *Eubacterium*, were positively correlated with most carbohydrate degradation pathways, such as starch degradation V, stachyose degradation, galactose degradation I and D-galactose degradation V (Fig. 3). These species are capable of fermentating general carbohydrates and producing butyrate, which are of anti-inflammatory effects in the gut (47, 48). Additionally, a few disease-enriched pathways previously linked to CRC were also found to correlate with disease-enriched microbial markers. For example, *Lachnospiraceae* bacterium 7 1 58FAA had a evident link with L-glutamate degradation V, a CRC-specific pathway via D-2 hydroxyglutarate that could drive epithelial-mesenchymal transition and induce CRC progression (49, 50). Similarly, some IBD-depleted species, such as *Alistipes shahii, Subdoligranulum spp*., and *Ruminococcus bromii*, had a negative association with superpathway of purine deoxyribonucleosides degradation, a pathway used as the source of energy (51) (Fig. 3). Of note, the pathways relating to amino acid degradation are positively associated with most of CRC-enriched bacteria, while negatively associated with IBD-enriched bacteria. Thus, clustered marker microbes could signify (at least in part) the changes in the overall metabolic capabilities in diseases and controls; in addition, these correlations between bacteria and microbial functions across studies and diseases revealed differences in metabolism among patients with different diseases, particularly between CRC and IBD.

**Fig. 3.**
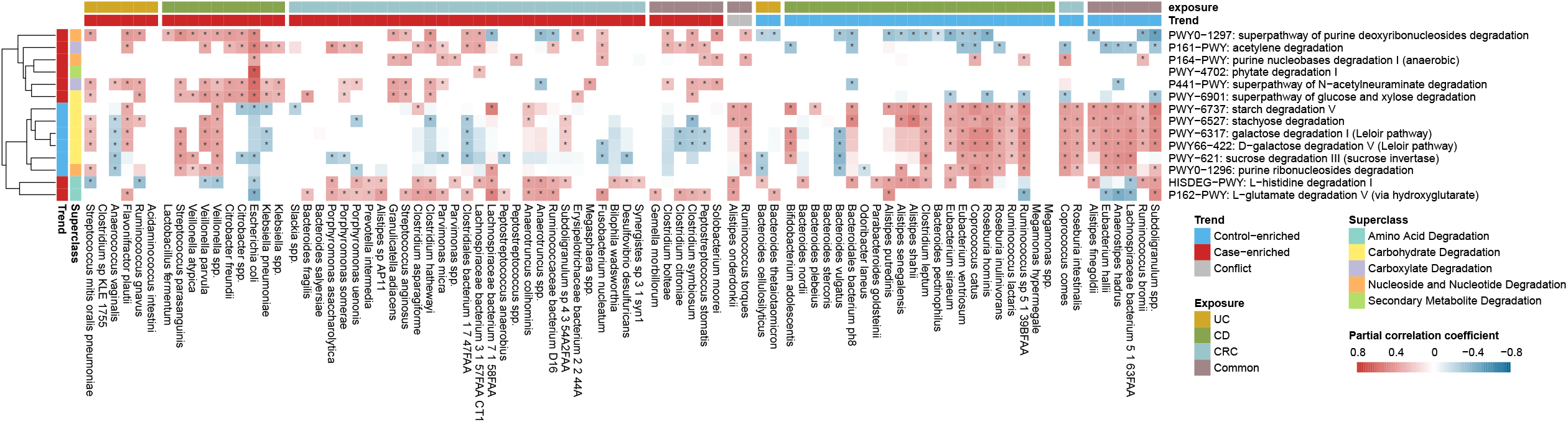
Marker microbes signified distinct degradation preferences. Shown here are the correlations of meta-analysis in relative abundances between the degredation pathways and the marker species (see Materials and Methods). The pathways were clustered using ‘mcquitty’ algorithm, while the species were sorted by their related diseases and changing trends. The blocks in the heatmap show the overall coefficients from the meta-analysis. The red block indicates positive correlation and blue indicates negative one. The asterisk indicates adjusted P-value of the overall coefficients in the meta-analysis is below 0.05. The similar plot for other pathways is shown in Fig. S5.

The gut metabolic properties are known to be influenced by food and microbial activities. Recenlty, researchers also revealed that cells/metabolites derived from the human host, likely due to compromised intestinal barrier (CIB), can also influence the growth of individual bacterium and the gut microbes as a whole (25, 28). CIB could lead to increased HDCs in the gut metagenomics. As expected, we found that HDCs were significantly elevated in cases of all datasets except PRJEB1220 (Table S1). Suprisingly, we found HDCs were also significantly correlated with some CRC-enriched species and half of control-enriched marker species in CD (Fig. S6). *Eubacterium ventriosum*, the control-enriched bacterial marker in CD datasets, was negatively correlated with HDCs in UC and CD (rho = -0.32, P-value = 8.16e-12 and rho = -0.25, P-value = 2.81e-6 respectively, Spearman’s rank correlation); the correlation was not significant in CRC, most likely due to its low abundances. *E. ventriosum* was previously shown to negatively correlate with fundamental components of eukaryotic cell membranes (25). Only three control-enriched species in UC had associations with HDCs (Fig. S6A), while most of UC-depleted pathways correlated with HDCs (Fig. S6B), implying that the metabolic functions had a better response to the intestinal status.

Together, our results revealed correlated changes between marker species and metabolic pathways, and suggested that both species and metabolic functions could be driven by the increased human derived contents leaked into the gut due to CIB, consisntent with our previous results (35).

### Marker species showed increased connectivity in diseases, presumably due to more stressed conditions

Having shown that the alterations in intestinal ecosystems could contribute to gut microbiota dysbiosis, we further explored the inter-relationships among the marker species within each physical condition. As ecologically important patterns, coexistence relationships within a biological community could reflect interplays between organisms and ecological roles of individual members. Applying co-occurrences analyses to gut microbes could help us compare coexistence patterns from different intestinal states, identify key species important to human health and provide an insight into maintenances of gut microbial ecosystems (52, 53).

We thus constructed inter-species networks using the disease marker species separately for cases and controls for each disease based on pairwise correlations of the species abundances. We used SparCC, a correlation method for microbiome data, to calculate the correlation coefficients among species to perform meta-analysis. We found that species in the cases were connected more often than they were in the controls of respective disease datasets (Fig. 4). For example, we found 172 positive pairs and 45 negative pairs of correlated marker species (fdr in meta-analysis < 0.05) in CRC, increased from 138 and 36 in the controls relatively (Fig. 4A & B, Fig. S7A &B). Similarly, we found much more positive and negative correlations among markers in cases than in controls in both CD and UC (Fig. 4A & B, Fig. S7C-F). These results were coincide with a previous study, which identified that the CRC patients networks contained more links among nodes than control networks, and the negative correlations declined when CRC patients under the chemotherapy (54).

**Fig. 4.**
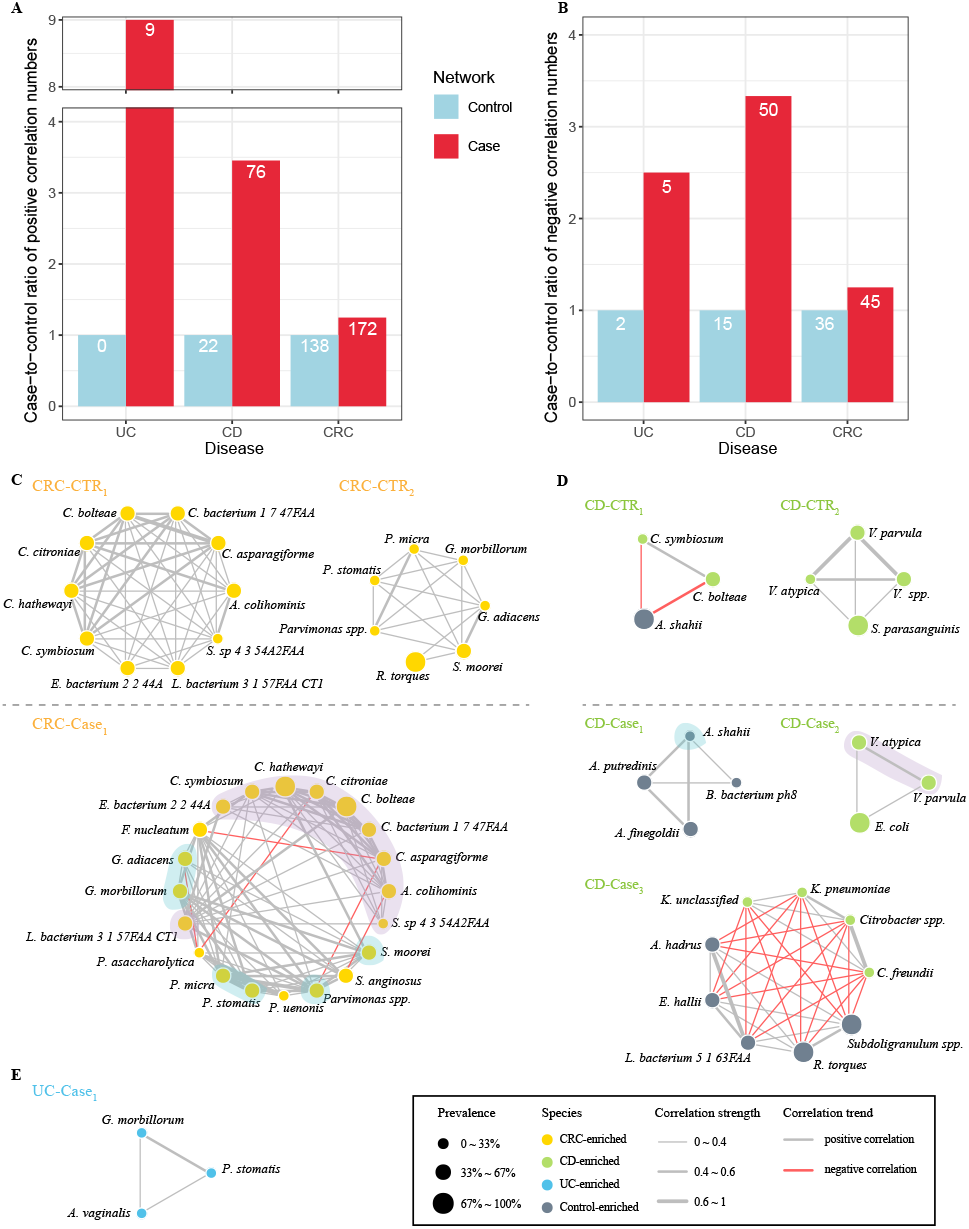
Increased correlations of marker species in diseased conditions. **A-B.**The statistics on the increases of positive correlated pairs **(A)** and negative correlated pairs **(B)** in case network (red) as compared with control network (blue) in each of the three diseases. Y-axis shows the case-to-control ratios of each disease, with number of pairs in the control network being normalized to 1. The numbers in the bars indicate the actual numbers of correlated pairs. **C-E.** Detected modules from correlation networks among marker species of per state in corresponding disease datasets (see Materials and Methods). Color of nodes means the alteration trends of species, and the sizes mean the prevalence of the bacteria in the overall health (or cases) of given datasets. Gray edges indicate positive relationship and red edges indicate negative. Thickness of edges indicates correlation strength.

In view of the difficulties to compare the networks as a whole, we used the mcode implemented in Cytoscape to detect modules, which are regarded as the highly interconnected clusters in a network and often used to gain biological insights from networks. In CRC datasets, we found that the modules from cases (named as ‘CRC-Case1’) had more members and was interconnected tighter as compared with those from controls (named as ‘CRC-CTR1’ and ‘CRC-CTR2’), (Fig. 4C). The species belonging to the same genus were associated more closely with each other, probably owing to their similar metabolic properties. Our network-derived modules could reveal previous known positive interactions. For example, *F. nucleatum*, a widely studied oral-associated anaerobe known to co-aggregate with other anaerobes to form biofilm and involve in the intestinal tumorigenesis (55, 56), showed positive correlations with *P. micra* in our case module (Fig. 4C) (57). Besides, though it lacks experimental evidences for our identified negative associations underlying CRC-enriched microbes, there were plenty of investigations about the competitive relationship among taxa during growth of biofilm (57-60). For example, *Porphyromonas gingivalis*, another known biofilm-forming partner of *F. nucleatum*, showed a negative correlation with *P. micra* (57). Thus, our results found the novel relations between CRC-enriched microbes and remained to be confirmed in further experiments.

Similar with CRC modules, modules from CD datasets also displayed the tighter relationships within the species at same taxonomic level, including members of genus *Klebsiella, Veillonella* and *Alistipes* (Fig. 4D). A previous study found that *Klebsiella* correlated positively with fecal calprotectin (FCP), an inflammatory marker for IBD, whereas *Ruminococcus* correlated negatively with FCP (53). In UC datasets, only the module from UC patients’ network (named as ‘UC-Case1’) was recognized (Fig. 4E). The strong correlation between *G. morbillorum* and *P. stomatis* was shown in UC module and CRC modules despite the sources of data, indicating the co-aggregation between them.

We also identified potential hub species in the networks using eigenvector centrality scores (ECSs) and betweenness centrality scores (BCSs) (Fig. S8). ECS served as assessment of node influence in a weighted network, measures the importance of the given node considering not only its connections with others, but also the connections of its related nodes. BCS was used to evaluate the transmission capacity of species. In CD datasets, we found that *Alistipes putredinis, A. shahii* and three CD-enriched *Veillonella* were the pivotal species in controls; while in CD patients, two CD-enriched Enterobacteriacceae, *Citrobacter spp*. and *Klebsiella* spp. took leading roles (Fig. S8C &D), both of which could deliver virulence proteins into host cells for assisting themselves against host immune system and infect mucosa so that thrive in gut (61-63). Nevertheless, health-related species were also the hub bacteria in patients with CD. These results support the view that we should be cautious to use antibiotic in CD therapy, which may disrupt fragile connections among species exhausting in cases, and cause some bacteria failure of recovery (64). As expected, CRC-enriched microbes, such as most of species from genus *Clostridium*, were at the center in both control network and patient network of CRC datasets (Fig. S8A &B). Although the top nodes with high BCSs were CRC-enriched species in both CRC control-network and CRC case-network, their niches changed, suggesting that the CRC-enriched microbes were vital in the dynamic network. In UC datasets, nodes with highest ECSs in controls network were UC-depleted species, while in the case network were mainly UC-enriched species (Fig. S8E &F). We found that the crucial nodes were also the members of corresponding module, validating representativeness of the modules in network.

Together, our inter-species network analysis revealed that marker species were more closely connected in diseased conditions; we speculated that due to oxidative stress and increased permeability of intestinal barrier, the gut ecosystem under diseased states may represent more stressful conditions, in which the growth of all microbes, especially the marker species, would be under stronger constraints and selection (11, 65). In addition, the hub species that were at the center positions and connect more with others, are more likely to be targets for disease treatment.

### Multi-class machine-learning models for disease stratification using gut microbial and metabolic markers

We next built multi-class models capable of distinguishing multiple diseases using the microbial and pathway profiles as well as those of the identified markers.

We first built four-class models with multiple types of features mentioned above using 10-time 10 folds cross-validation (see Materials and Methods). A model based on combined taxonomic and metabolic profiles (all features) reached a highest accuracy with true positive rate (TPR) of 0.8 (Fig. S9A &B). We achieved significantly better classification performance on UC and CD than a recent study, with TPRs of 0.81 and 0.91 for UC and CD samples respectively in this model (Fig. S9B), as compared to 0.51 and 0.67 in the Fig. 6 of literature (25) where the model was also trained on combined profile. Models built on either taxonomic (TPR as 0.69) or functional profiles (0.74) showed decreased performance (Fig. S9A). The classifier based on combined markers also performed well with an overall accuracy of 0.75 (Fig. 5B). The classification errors were mostly contributed by UC samples, which associated with the least number of disease markers and the least proportion of unique ones (Fig. 1A &Fig. S4). Furthermore, UC shared majority of its markers with CD, but not *vice versa;* consequently, a significant proportion of UC samples were classified as CD, but only a few of CD samples were misclassified as UC (Fig. 5B).

**Fig. 5.**
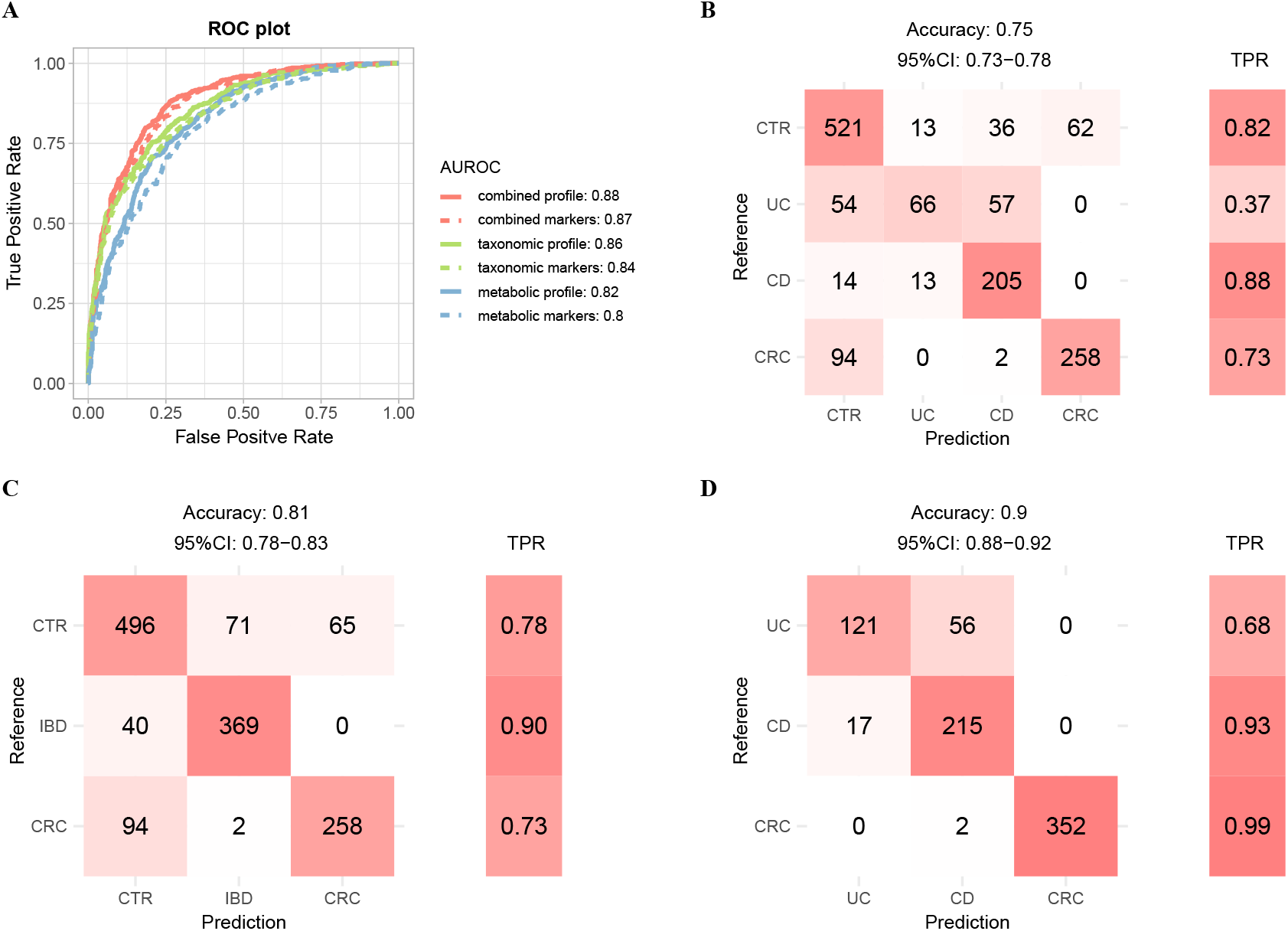
Random forest models for patient stratification using taxonomic and/or metabolic profiles. **A.** Binary models for distinguishing cases from controls were constructed using various type of features as shown in plot. The classification results were ploted as the AUROCs of corresponding model. AUROC is evaluated through ten-time ten-fold random forest cross-validation in all samples (see Materials and Methods). The combined profile indicated the relative abundance profile of all taxonomic and metabolic features. The combined markers indicated the relative abundance of all taxonomic and metabolic markers. **B-D.** Confusion matrix evaluation of the four-class model (**B**), three-class model (**C**), and cases model (**D**) based on combined marker features for distinguishing different physical condition. The number in row i and column j in the matrix on the left part indicates how many patients with disease i actually were categorized to disease j in model. Color filling the cell means the relative size of the number in the corresponding row. The right part is the TPR for per disease type. Total accuracy indicates the fraction of all correct predictions, and 95% CI is the confidence interval of accuracy.

Since UC and CD are sub-types of IBD, we thus combined their samples into the IBD group and built three-class models (i.e., IBD, CRC and controls, Fig. 5C &Fig. S9A); as shown in Fig. 5C, we achieved much better classification rate with an overall TPR of 0.81 using also the combined markers. 90% of the IBD samples were classified correctly; the remaining 10% were misclassified as control, but none was classified as CRC.

We also built two additional models, including a “case-control” model to distinguish cases from controls (also referred to as “binary” model), and a “cases” model to assign cases to distinct diseased states (also referred to as “cases” model). The cases model was particularly important and clinically relevant due to the clinical overlap in presentation of these diseases, as well as the risk of IBD patients to eventually develop CRC. We evaluated the performance for the binary models on the 632 controls and 763 cases and found that all models performed well, with the area under receiver operating characteristic curves (AUROCs) ranging from 0.80 to 0.88. Notably, taxonomic-based models in generally performed better than the metabolic-based models, as the model with the combined profile attained a highest accuracy (Fig. 5A). Surprisingly but expected, models using only the marker species/pathways performed comparable to those that used all species/pathways, especially the combined-based model, suggesting that the much-shortened list of markers are of practical and clinical value. For the “cases” models, the classifier based on combined profile achieved an accuracy of 0.98, while the accuracy of classifier based on combined markers only achieved 0.9. The metabolic-based model still performed better than taxonomic-based model, indicating the functions of microbes reflected the gut status better than species distribution (Fig. 5D, Fig. S9A). We noticed significant differences among diseases in terms of TPRs in the “cases” models. In particular, we achieved high accuracies for CRC and CD (TPR as 99% and 93% respectively), as compared with the relatively low TPR for UC (68%). The latter was likely due to the fact that UC shared most of its markers with CD and had only few unique markers; consequently, most of the misclassified UC cases were predicted as CD (Fig. 1A &Fig. S4).

To evaluate if the performances were not biased by a single dataset, we applied leave-one-dataset-out (LODO) analysis, which left one dataset as the testing data, and utilized the remained datasets to train the random forest models. The LODO models based on combined profile and combined markers (referred to as “Four-class All” and “Four-class Dif” relatively) to distinguish cases with the different disease and controls, achieved an average accuracy of 0.81 and 0.75 on training data respectively, and an overall accuracy of 0.56 and 0.65 on testing data respectively (Fig. S9C). The training results of each dataset were similar, indicating there was no bias across datasets. Moreover, except the models for classifying patients with different disease, the models trained on the combined markers performed better than the corresponding models trained on combined profile.

These results suggest the classifiers based on combined markers could achieve similar accuracies with those based on combined profile, indicating the clinical feasibility of the microbial markers. Besides, additional information other than fecal metagenomics, such as physiological, genetic and clinical information of the human hosts are required to further improve the prediction accuracies.

## DISCUSSION

In this study, we collected fecal metagenomics datasets for three common intestinal diseases, namely CRC, CD and UC, totaling 11 projects, 13 datasets, 763 patients and 632 controls. We selected these diseases because they all have strong associations with gut microbiota dysbiosis, share clinical presentations and are pathogenically linked, i.e. both UC and CD patients are at high risk of developing CRC. We performed meta-analysis and identified in total 87 marker species and 65 marker pathways that were consistently changed (i.e. case-depleted or case-enriched) in the same disease. We grouped the marker species into disease-specific and common clusters according to whether or not the member species were unique to a certain disease, and revealed their distinct phylogenetic relationships: for example, CRC-specific species are more diverse phylogenetically than UC- and CD-specific markers. Strikingly, UC- and CRC-specific species are phylogenetically closer to each other than to those of CD (Fig. S3), in part due to the fact that UC-patients are at higher risk of developing CRC (9).

We then characterized the marker pathways. We first revealed that each disease formed their exclusive module profiles, that the CRC patients had an elevated trend in amino acid degradation while the CD patients behaved in an opposite way. We then explored that almost all marker pathways correlated significantly with marker species (Fig. 3 &Fig. S5); additionally, clustered marker species tended to correlate significantly with the same sets of pathways. These results were not unexpected since marker species that are closer phylogenetically tend to have similar metabolic capacities. We then noticed strong correlations between a significant proportion of marker species and HDCs. HDC has been shown to be significantly increased in many intestinal diseases, and could be used as an indicator for the extent of leaky gut caused by CIB (Table S1). The elevated HDCs in all datasets may signify significant changes in physio-metabolic properties of the local gut environment due to leaked human derived contents under diseased states. Our results thus suggested that human derived contents due to CIB could have stronger impact on gut microbiota than we have previously anticipated. Finally, by considering the gut microbiota as an ecosystem, we revealed that marker species showed increased connectivity in diseases as compared with the respective controls, and control-enriched species together with pathogens played important roles in ecological network of CD patients. Thus we speculate that the diseased gut may represent a more stressful environment due to the physio-metabolic changes including oxidative stress and/or bleeding; the inhabitant microbes are thus under stronger selection, and show either more cooperation (positive correlation) or competition (negative correlation). And our results support the view that we should be cautious to use antibiotics in therapies for CD patients.

Utilizing the identified marker species and pathways, we obtained four high-performance models for disease identification and patent stratification. The first “four-class” model could separate samples into controls or individual diseases (Fig. 5B), with an overall TPR of 0.75. UC has the lowest TPR (0.37, Fig. 5B) in this model; however, most of the wrongly classified samples went to CD, consistent with previous efforts (25) and the fact that UC had very few unique markers and shared most of its marker with CD (Fig. 1A and Fig. S4). Regardless, it represents one of the best models that could classify IBD subtypes with TPR as 0.81 and 0.91 separately in UC and CD samples, while in a previous study TPR from the cross-validation model built on the abundance of metabolites and species only achieved 0.49 and 0.66 in UC and CD respectively (25). We also built three additional models, including a “binary” model to distinguish cases from controls, a “three-class” model to consider the IBD as a whole and distinguish patients with cancer or inflammation from controls, and a “cases” model to assign cases to distinct diseased states. In our opinion, both are relevant in clinical applications. For example, the binary model, with AUROC of 0.87, can inform the subjects for further clinical inspections such as colonoscopy, while the “three-class” model, with an overall TPR of 0.81, can evaluate the patients for potential IBD and CRC risks. The “cases” model with a high accuracy of 0.9 was worth watching, due to the clinical overlap in symptoms of these intestinal diseases, as well as the risk of IBD patients to eventually develop CRC.

Taken together, our results demonstrated the necessity and feasibility of metagenome-based multi-disease classifications. The few selected marker species and pathways had similar performances to all the taxonomic and metabolic features, and could be easily translated to clinical uses. Our meta-analysis methods and cross-disease comparisons improved our understanding on the differences and relationships among common intestinal diseases that could have similar clinical symptoms; and could be expanded to include more gastrointestinal disorders such as irritable bowel syndrome and colon polyps.

## MATERIALS AND METHODS

### Data collection and pre-processing

We obtained in total 175 records by searching public metagenomic databases including NCBI PubMed (66) and GMrepo (67) using key words such as metagenomics and relevant disease names (see Fig. S1 for details). We aimed to collect metagenomics sequencing data with high-resolution for better understanding the functions of microbes. After filtering out the duplicates, 16s rRNA sequencing data and the metagenomics data without detailed metadata or not meeting minimum samples requirements, we selected in total thirteen metagenomics datasets, including three, three, and seven datasets for CD, UC and CRC respectively. Please consult Fig. S1 for the selection procedure and results, and Table S1 for the thirteen datasets.

Raw sequencing reads were retrieved from European Nucleotide Archive (ENA) (68) under the following identifiers: PRJEB6070 (22), PRJEB27928 (33), PRJEB12449 (69), PRJEB10878 (26), PRJEB7774 (39), PRJDB4176 (33), cohort 1 of PRJNA447983 (33), SRP057027 (24), PRJEB1220 (70), PRJNA400072 (25) and PRJNA389280 (71); sample metadata were also downloaded from ENA. For projects containing samples resulting from longitudinal surveys, i.e., participants were sampled multiple times over extended periods of time and/or during treatment/intervention, including SRP057027, PRJEB1220 and PRJNA389280, we selected the first time-point from each participant to avoid false positive in following analysis. In total, we obtained in total 632 non-disease controls and 763 patients for the following meta-analysis, including 354, 177 and 232 samples of CRC, UC and CD (Table S1).

### Taxonomic and functional profiling of metagenomics data

To keep only the high quality data, low quality reads and adapters were firstly removed via Trimmomatic (version 0.35) using the Truseq3 adapter files (TruSeq3-PE.fa for paired-end data and TruSeq3-SE.fa for single-end data) and a MINLEN cutoff of 50 (72). The remaining “clean” reads were then mapped to the human reference genome (hg19) using bowtie2 (version 2.3.4.3) (73) with default settings to identify and remove human reads. The identified human reads were also used to compute HDC for each sample as the percentage of mapped reads out of total clean reads, which have been shown to be a marker for intestinal barrier dysfunction and correlate with the marker species of several intestinal diseases (35). For samples that were sequenced multiple times (e.g., for the purpose of increasing sequencing depths), the resulting multiple sequencing files were merged before further analysis. The merged and clean non-human reads were then quantified in taxonomic and functional levels using MetaPhlAn2 mapping to mpa_v20_m200 database and HUMAnN2 mapping to ChocoPhlAn database and full UniRef90 database (74, 75).

To avoid the noise of low-abundance, pathways with zero value in over 15% samples of a dataset were excluded. Species and pathways that did not meet a maximum relative abundance cutoff of 1×10^−3^ and 1×10^−6^ separately in at least 50% datasets for a specified disease were removed. The abundance data were then loaded into R (ver 3.6.3 mainly; https://www.r-project.org) and analyzed.

### Controlling for confounding factors and identification of marker species and pathways

Within-project confounding factors, i.e., those showed significant differences between phenotype groups in a dataset were firstly identified using Wilcoxon rank sum test or chi-squared test on per-dataset basis. Then, the identified confounding factors (see Table S1 for the results) in differential analysis were controlled for using MaAsLin2 package in R ver 4.0.0, a multivariable analysis tool to adjust the covariates and identify association effects of the species and pathways to disease in each dataset. Accounting for the heterogeneity between datasets, we performed meta-analysis to aggregate the association effects via MMUPHin package in R ver 4.0.0, and identify the final “maker” species and pathways. Here an adjusted p-value < 0.05 from meta-analysis was used as the cutoff for the makers.

### Clustering of disease-enriched species and their prevalence in the three diseases

Consistently disease-enriched marker species (i.e., those were marker species in at least two datasets of the same disease) were grouped into disease-specific or common to multiple diseases according to their association with the diseases (Fig. 1A). To observe the prevalence of the clusters in the overall patients and a unique disease (i.e., CRC, CD and UC), their prevalence in the diseased samples were first calculated: for each selected marker species, its 95% percentile abundance in all controls was used as a cutoff to defined its presence (“1”, i.e., its relative abundance in the sample was higher than the 95% quantile relative abundance of all control samples) or absence (“0”). By doing so, we obtained a binarized matrix with each row representing a disease-enriched marker microbe, and each column representing a patient. The prevalence matrix from all patients was used to calculate the Jaccard distances among the species using the diversity function of vegan package. We compared the inner Jaccard similarities among the clusters of disease-enriched marker species using Wilcoxon rank sum test (for pairwise comparisons) and Kruskal-Wallis rank sum test (for multi-group comparisons). The prevalence of each cluster between patients and controls in each disease were also compared using the Cochran-Mantel-Haenszel test with “dataset” as the blocked object by cmh_test function of coin package.

### Phylogenetic relationship of disease-enriched marker species

To show the phylogenetic relationships among the disease-enriched marker species, a phylogenetic tree was generated based on their NCBI taxonomy using an online tool, phyloT (https://phylot.biobyte.de/), setting internal nodes collapsed and polytomy as no. The tree file then was visualized using Evolview ver3, a webserver for annotation and management phylogenetic trees (76). The nodes were colored depending on their corresponding clusters identified in previous section. The last common ancestors (LCAs) were determined according to the NCBI taxonomy of the species in corresponding branch.

### Identification of HDC-correlated features

For each dataset, Spearman’s rank correlation was used to identify HDC-related microbial features (e.g., species and functions) using a p-value cutoff of 0.05. Features that maintained a significant positive or negative relationship with HDC in at least two datasets of a disease were identified as HDC-related features.

### Functional profile of metabolic modules

According to the category of MetaCyc database, we concluded the microbial functions into their corresponding superclasses as metabolic modules (42). The expression of each metabolic module was summarized as the average logarithm relative abundance of its contained functions. Setting the quantiles from 0.1 to 0.9 and the increment as 0.1, we calculated the generalized fold changes of modules between the controls and cases, and performed the Wilcoxon rank sum test with the “datasets” as the blocked object to evaluate the differences.

### Microbial ecosystem analyses using species-species correlations

To characterize the relationships among the marker species and the resulting interaction networks, SparCC, a sparse correlation method for compositional data (77) was used to identify correlations among marker species. SparCC was previously shown to be able to reduce the high false positive by Spearman’s rank correlation in metagenomics data. The tool requires read counts as input, therefore we multiplied the relative abundances of the species to the number of reads mapping to mpa_v20_m200 database, and got the microbial counts of each sample. For each dataset, species-species correlations were calculated for control and case samples separately. By setting both the iteration number and simulation as 100 and threshold of correlation strength as 0.05, SparCC generated the correlation matrices of the real data and 100 simulated datasets. The pseudo P-values were assessed as the proportion of simulated datasets with a correlation value at least as extreme as that calculated from the real data. After filtering correlations with P-value < 0.05, we performed meta-analysis to aggregate correlation coefficients for each disease type via random-effects model, which summarizes overall correlation based on Fisher’s z transformation with metacor function (32, 78). The summarized correlations with adjusted p-values <0.05 in meta-analysis were used to construct networks. The networks were analyzed in Cytoscape (79) to identify modules using mcode with default parameters. We then evaluated eigenvector centrality and betweenness centrality of networks using correlation strength as weight, and visualized networks with igraph package in R. The size of node indicated prevalence of the bacteria in counterpart samples. Positive and negative correlation coefficients as strength of edges were painted gray and red separately.

### Correlating functional profiles with species

To identify species underlying functional changes in the metagenomic data, correlations between relative abundance values of marker species and marker metabolic pathways in each dataset were computed using partial Spearman’s rank correlation to adjust the identified covariates. The relative abundances were log-transformed; to avoid Inf values, pseudo values 1e-06 and 1e-09 respectively were added to taxonomic and functional abundances before log-transformation. The resulting correlations with P-value <0.05 were retained to perform meta-analysis and get the overall correlation coefficients with metacor function.

### Random forest classifiers and cross validation

To check if the metagenomics data could be used to distinguish different diseases from each other and/or from healthy controls, the random forest function of the randomForest package were used to build several machine-learning classifiers. Samples were split into training and test datasets during the modeling; to prevent biases due to one-time split, a ten-times and ten-fold cross validation technique was used by using the caret package. Thus, for each model, in total 100 models were created; the overall performance was the average of all the 100 models.

For the overall cross-validation, we pooled datasets into one, then applied logarithm transformation and standardization to the taxonomic and functional abundance profiles. The data were split into training set and testing set repeatedly 10 times. All models trained on the training set were applied to corresponding testing set and the prediction scores were averaged. For binary classifiers, i.e., classifiers that attempt to classify samples into two distinct groups: diseased or control, the AUROC values were used to evaluate their performance. For the multi-class classifiers, i.e., classifiers that attempt classify samples into multiple distinct groups such as CRC, UC, CD and control, the detailed predicting results were shown as confusion matrixes and the performances as overall accuracies. We also built models based on the identified microbial markers to test if the performances of models were improved.

LODO analysis was also performed to test if the cross-validation models were biased due to one specific dataset. In short, all but one datasets were pooled to build models with 10-fold and 10-times cross-validation as described earlier, and resulting model was then applied to the left-out dataset. To optimize each model after each split, we set the ranges for number of trees and number of features to tune the hyperparameters with the mlr3 package. The whole process was repeated several times until every dataset was used the left-out dataset in turn.

### Other statistical tests

We calculated alpha diversity of each dataset with the diversity function of vegan package. Two-sided Wilcoxon rank sum test was used to compare two sets of numeric data; the wilcox_test function implemented in the coin package was used to block with the factor “dataset”.

### Data availability

The datasets generated and R codes during this study are available at https://github.com/whchenlab/2019-puzi-multi-gut-disease-classifier. Correspondence and requests for materials should be addressed to W.H.C. and X.M.Z…

## Data Availability

The datasets generated and R codes during this study are available at https://github.com/whchenlab/2019-puzi-multi-gut-disease-classifier. Correspondence and requests for materials should be addressed to W.H.C. and X.M.Z...

## ACKNOWLEDGMENTS

We thank Na L Gao, Lei Liu and Xinming Li for valuable discussion.

This work was partly supported by National Key Research and Development Program of China [2019YFA0905600 to W.H.C.], National Natural Science Foundation of China [61932008, 61772368, 61572363 to X.M.Z], National Key R&D Program of China [2018YFC0910500 to X.M.Z], Natural Science Foundation of Shanghai [17ZR1445600 to X.M.Z], Shanghai Municipal Science and Technology Major Project [2018SHZDZX01 to X.M.Z] and ZJLab. The funders had no role in study design, data collection and interpretation, or the decision to submit the work for publication.

W.H.C. and X.M.Z. designed the study. P.J. and S.W. collected and analyzed the data, Q.L. coordinated the data downloads and analysis. W.H.C., and P.J. wrote the manuscript with all authors contributing to the writing and providing feedbacks. All authors read and approved the final version of the manuscript.

We declare no competing interests.

## Supplemental Material

**Table S1. A list of CRC and IBD datasets used in the study and statistical results of confounding factors, HDCs and Shannon diversity.** The project “SRP057027” lacks detailed information about age, BMI and gender, but the original literature gave a statistical table which shows that there were no significant differences in age and gender between controls and CD patients (24). Numerical variables were reported as the median values in the corresponding group. The P-values were calculated using Wilcoxon rank sum test or chi-squared test to compare cases to controls.

**Fig. S1. The pipeline for metagenomics data collection.** We collected records about IBD and CRC in public databases until Sep, 2019. The requirements for samples were that (1) the number of each group in each dataset was not below 20, and (2) there were no overlapping samples across datasets.

**Fig. S2. The associated effects of identified marker species in each dataset of each disease.** The marker species which were also significantly differential species in corresponding dataset (P-value < 0.05) were plot. Each block indicates a marker species of corresponding disease. Red means that the case-enriched species and blue means the control-enriched one. **A**, CRC; **B**, CD; **C**, UC.

**Fig. S3. Phylogenetic relationships of disease-specific and shared marker species.** Phylogenetic relationships of the marker species were based on the NCBI common tree, generated using the online phyloT tool (https://phylot.biobyte.de), and visualized using Evolview ver3.0 (76) (with manual annotations). The LCAs were determined according to the NCBI taxonomy information of the species in corresponding branch.

**Fig. S4. Identified marker pathways that showed consistent changes in the respective diseases.** The relative abundances of pathways were identified using HuManN2 on the metagenomics data. Pathways significantly enriched in cases (or controls) of corresponding disease in meta-analysis are shown (fdr < 0.05, Benjamini-Hochberg FDR correction, see Materials and Methods). Red block indicates case-enriched pathway, and blue block indicates control-enriched one.

**Fig. S5. Marker microbes showed distinct biosynthesis and other metabolic pathways preferences.** Similar to Fig. 3, however, shown here are the correlations of meta-analysis in relative abundances between the non-degradation pathways and the marker species (see Materials and Methods). The pathways were clustered using ‘mcquitty’ algorithm, while the species were sorted by their related diseases and changing trends. The blocks in the heatmap show the overall coefficients from the meta-analysis. The red block indicates positive correlation and blue indicates negative one. The asterisk indicates adjusted P-value of the coefficient in meta-analysis is below 0.05.

**Fig. S6. Some marker species and pathways correlated significantly with HDCs.** Each block indicates the marker species (**A**) and marker species (**B**) identified in meta-analysis. Red means that the species/pathways was HDC-related evidently in at least two datasets of corresponding disease type. Blue means the species/pathways was not HDC-related.

**Fig. S7. Correlation networks among disease-altered species in CRC datasets (A-B), CD datasets (C-D) and UC datasets (E-F).** Color of nodes means the alteration trends of species in its corresponding datasets. The sizes mean the prevalence of the bacteria in the overall controls (or cases) of corresponding datasets. Gray edges indicate positive relationship and red edges indicate negative. Thickness of edges indicates correlation strength.

**Fig. S8. Evaluation for ECSs and BCSs of networks showed in Fig. S7.** Blue bars indicate the centralies of control network from datasets, and red bars indicate centrailies in case network. **A-B**, CRC; **C-D**, CD; **E-F**, UC.

**Fig. S9. Cross-validation models (A-B) and LODO models (C) based on various types of features for different purposes. A:** The pheatmap despicts the cross-validation results of multi-class models built on corresponding features. Rownames indicate the classification tasks (Four-class: CTR/UC/CD/CRC; Three-class: CTR/IBD/CRC; Cases: UC/CD/CRC). Column names indicate the type of features. Each block indicates the accuarcy of each model. **B:** Confusion matrix evaluation of four-class model built on combined profile in taxonomic and functional level for distinguishing different disease and controls. The number in row i and column j in the matrix on the left part indicates how many samples with state i actually were categorized to state j in model. Color filling the cell means the relative size of the number in the corresponding row. The right part is the TPR for per physical condition. Total accuracy indicates the fraction of all correct predictions, and 95% CI is the confidence interval of accuracy. **C.** The pheatmap despicts the training results and validation result of each LODO model. Each row indicates the result of the LODO models training in the remainig data except the appointed dataset (see Materials and Methods). Column names were the combination of the classification task and type of features. For example, the ‘Four-class All’ means the LODO models based on combined profile were used to distinguish four classes (CTR/UC/CD/CRC), when the ‘Four-class Dif’ means means the LODO models based on combined markers to distinguish four classes. In this plot we only showed the models based on either combined profile (All) or combined markers (Dif). For the multi-class models, including the models starting with ‘Four-class’, ‘Three-class’ and ‘Cases’, we used the accuracy as the evaluation of classification. For the binary models, including ‘Binary All’ and ‘Binary Dif’, were displayed as AUROCs. The model average were the mean values of each column. The LODO validation were the integrated validation results of the LODO models in the same column.

