## Supplementary material for "Metagenomic analysis of common intestinal diseases reveals relationships among microbial signatures and powers multi-disease diagnostic models": Fig. S9

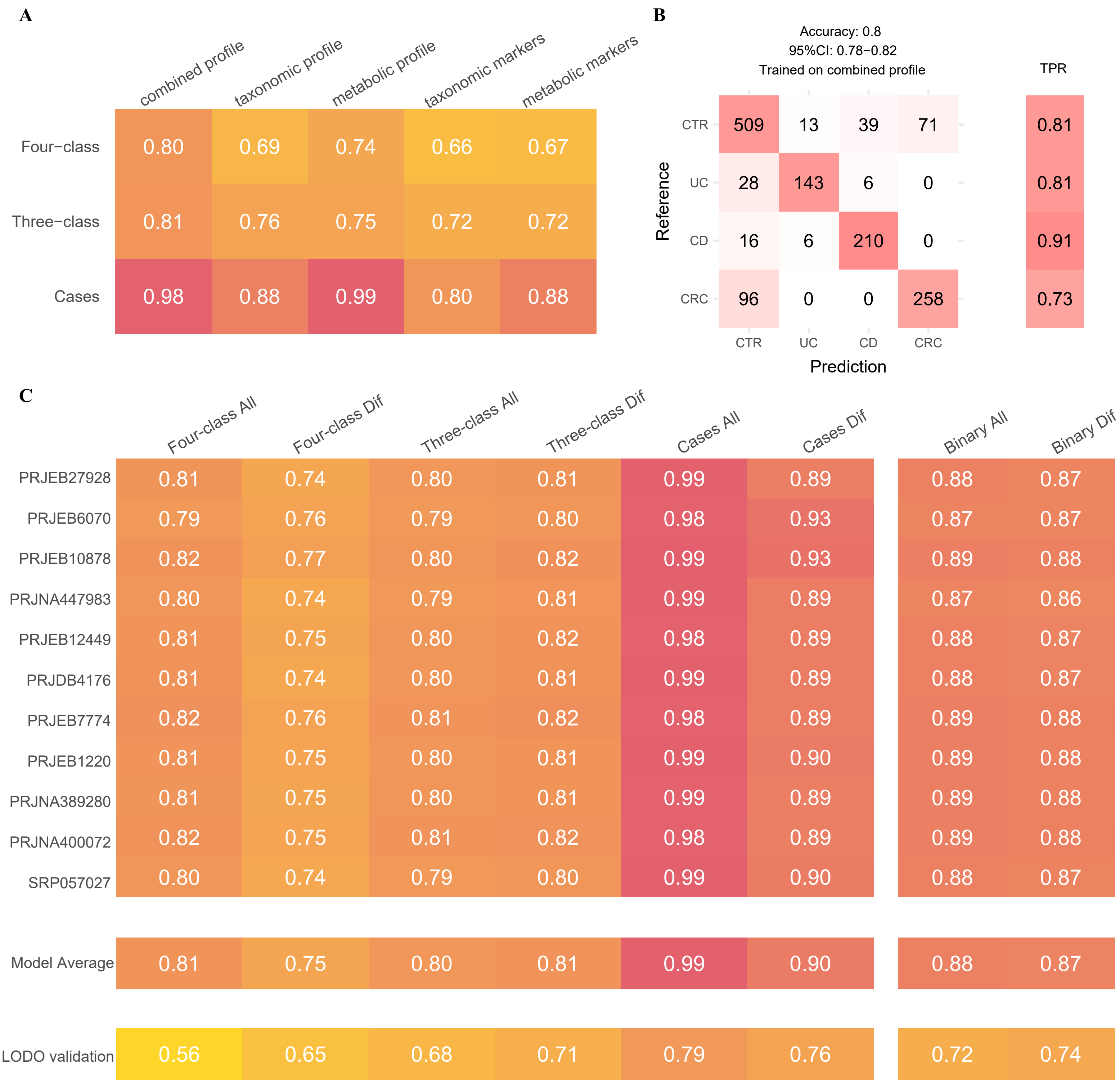

A

| | combined profile | taxonomic profile | metabolic profile | taxonomic markers | metabolic markers |
|---|---|---|---|---|---|
| Four−class | 0.80 | 0.69 | 0.74 | 0.66 | 0.67 |
| Three−class | 0.81 | 0.76 | 0.75 | 0.72 | 0.72 |
| Cases | 0.98 | 0.88 | 0.99 | 0.80 | 0.88 |

B

Accuracy: 0.8
95%CI: 0.78−0.82
Trained on combined profile

| Reference \ Prediction | CTR | UC | CD | CRC | TPR |
|---|---|---|---|---|---|
| CTR | 509 | 13 | 39 | 71 | 0.81 |
| UC | 28 | 143 | 6 | 0 | 0.81 |
| CD | 16 | 6 | 210 | 0 | 0.91 |
| CRC | 96 | 0 | 0 | 258 | 0.73 |

C

| | Four-class All | Four-class Dif | Three-class All | Three-class Dif | Cases All | Cases Dif | Binary All | Binary Dif |
|---|---|---|---|---|---|---|---|---|
| PRJEB27928 | 0.81 | 0.74 | 0.80 | 0.81 | 0.99 | 0.89 | 0.88 | 0.87 |
| PRJEB6070 | 0.79 | 0.76 | 0.79 | 0.80 | 0.98 | 0.93 | 0.87 | 0.87 |
| PRJEB10878 | 0.82 | 0.77 | 0.80 | 0.82 | 0.99 | 0.93 | 0.89 | 0.88 |
| PRJNA447983 | 0.80 | 0.74 | 0.79 | 0.81 | 0.99 | 0.89 | 0.87 | 0.86 |
| PRJEB12449 | 0.81 | 0.75 | 0.80 | 0.82 | 0.98 | 0.89 | 0.88 | 0.87 |
| PRJDB4176 | 0.81 | 0.74 | 0.80 | 0.81 | 0.99 | 0.89 | 0.88 | 0.87 |
| PRJEB7774 | 0.82 | 0.76 | 0.81 | 0.82 | 0.98 | 0.89 | 0.89 | 0.88 |
| PRJEB1220 | 0.81 | 0.75 | 0.80 | 0.81 | 0.99 | 0.90 | 0.89 | 0.88 |
| PRJNA389280 | 0.81 | 0.75 | 0.80 | 0.81 | 0.99 | 0.89 | 0.89 | 0.88 |
| PRJNA400072 | 0.82 | 0.75 | 0.81 | 0.82 | 0.98 | 0.89 | 0.89 | 0.88 |
| SRP057027 | 0.80 | 0.74 | 0.79 | 0.80 | 0.99 | 0.90 | 0.88 | 0.87 |
| Model Average | 0.81 | 0.75 | 0.80 | 0.81 | 0.99 | 0.90 | 0.88 | 0.87 |
| LODO validation | 0.56 | 0.65 | 0.68 | 0.71 | 0.79 | 0.76 | 0.72 | 0.74 |
